# Applying AI models to digital placental photographs to automate and improve morphology assessments

**DOI:** 10.64898/2026.02.28.26347346

**Authors:** Alison D Gernand, Rachel E Walker, Yimu Pan, Manas Mehta, Gwen Sincerbeaux, Kelly Gallagher, Lisa M. Bebell, Joseph Ngonzi, Janet M. Catov, Lauren B. Skvarca, James Z Wang, Jeffery A Goldstein

## Abstract

**Background:** Placental growth and function are imperative for healthy fetal growth; data on placentas can inform research and clinical care. Measuring placental size after delivery should be easy, but current methods are hard to standardize and error prone. We developed PlacentaVision using artificial intelligence (AI)-based models, to automatically, accurately, and precisely measure placentas from digital photographs.

**Objective:** We aimed to compare placental disc morphology between gross pathology examination (human measurements) and our automated PlacentaVision model (AI measurements).

**Methods:** PlacentaVision is a multi-site study to assess placental morphology, features, and pathologies from digital photographs. We built a large dataset of digital placenta photographs and clinical data from singleton births at three large hospitals: Northwestern Memorial (Chicago; n=24,933), UPMC Magee-Womens (Pittsburgh; n=1198) and Mbarara Regional Referral (Uganda, n=1715). Data and images were from the medical record for Northwestern, part of a biobank study for Magee, and from our prospective studies for Mbarara. We compared long and short disc axis length (defined by Amsterdam criteria) between human and AI-based PlacentaVision measurements by calculating the difference and using Bland-Altman; we stratified by site, disc shape, infant sex, and term/preterm birth.

**Results:** Mean (SD) disc length was 19.2 (3.1) and 18.6 (3.1) cm from PlacentaVision and human measurement, respectively, with a difference of 0.57 (2.19) cm. Disc width was 16.3 (2.3) cm and 16.1 (2.4) cm from PlacentaVision and human measurement, respectively, with a difference of 0.25 (1.85) cm. Bland-Altman limits of agreement were −3.7 to 4.9 cm for length and −3.4 to 3.9 cm for width. Irregularly-shaped placentas had a greater difference between PlacentaVision and human measurements compared to those with round/oval shapes (length differences of 1.53 and 0.45 cm respectively). Further, there were length differences by site (Northwestern 0.6, Magee 0.0, and Mbarara 0.4) and gestational age at birth (preterm 0.71, term 0.53 cm), but similar results for male and female placentas. Results for width were similar to length.

**Conclusions:** AI-based measurements were less than a cm from human measurements overall. Our findings of larger differences for irregular shapes and preterm may indicate it is difficult for humans to measure irregular or small placentas according to protocol. PlacentaVision can automate and standardize the process.

## Introduction

The human placenta is invaluable in prenatal development and future human health outcomes, but it is often overlooked in both biomedical research^1^ and clinical care.^2,3^ Over three decades ago, during the early development of the Barker hypothesis which eventually became the Developmental Origins of Health and Disease (DOHaD) hypothesis, David Barker and colleagues found that placental weight and adult blood pressure were positively associated.^4^ Furthermore, blood pressure during adulthood was highest in those with a small birth weight but large placenta. Subsequent work continued to link placental size to health outcomes.^5,6^

Studies associate placental size, surface area and weight with birth outcomes including infant size^7–9^ and adverse perinatal oucomes.^10^ Childhood blood pressure at 4 and 9 years old has been associated with placental weight^11^ and size and shape of the placenta surface.^12^ Long term offspring health outcomes such as hypertensive disorder and other cardiac related diseases,^13–18^ asthma,^19^ various cancers,^20–22^ overweight,^23^ mental health disorders,^24,25^ and mortality – all cause^26^ and cardiac related^27,28^ – have all been associated with aspects of placental size, shape, area or thickness. This research consistently demonstrates that variations in placental morphology – particularly length, width and area are associated with important health outcomes.

Placental pathology has consistently been impeded by the limited number of placental pathologists and differences in systems and classifications.^29^ Historically, protocols for measuring and diagnosing placentas have varied across hospitals, laboratories and studies,^30^ spurring experts to convene the Amsterdam Placental Workshop and define sampling and lesions in 2014.^31^ However, while the need for further standardization remains, automation is the new frontier to advance placental assessment in a meaningful way. Uniform protocols are helpful, but humans still interpret these in different ways. Additionally, only about 20% of placentas are assessed in the US, in part due to the high resource burden of pathology. Artificial intelligence (AI)-based models could be the answer to standardization *and* automation that could allow many more (if not all) placentas to be assessed.

One of the most promising solutions to human measurement variation is automation using digital images.^32^ Pathologists recommend photographs to be part of a standard exam,^33,34^ and some have recognized the potential for more precise measurements from photographs.^34^ The PlacentaVision project was conceived to develop a rapid, accessible, and cost-effective AI-driven tool that analyzes photographs of placentas taken at birth to detect abnormalities and assess potential health risks. The key components of the system include morphological characterization through robust semantic segmentation,^35–37^ and the identification of placental features and diagnoses using pathology reports as training targets.^38–40^ This tool can read a ruler in the image to calculate the long and short axes, classify shape, and estimate the weight of the disc. How these AI measurements compare with humans has not yet been quantified, and in contrast to lesions diagnosed histologically by pathologists, human gross measurements are not ground truth.

In this study, we aimed to compare placental disc morphology between human measurements during gross pathology examination and automated PlacentaVision measurements from digital photographs. We hypothesized that measurements would disagree to some extent due to the inconsistency of human measurements in a gross exam and the lack of clarity in some pathology protocols and definitions. As well, we expected error in our automated models (e.g. seeing membranes as the disc). We expected closer agreement with regularly shaped placentas (round/oval) compared to irregularly shaped placentas but examined differences by hospital, infant sex, and gestational age at birth without a directional hypothesis.

## Methods

We conducted a comparison study^41^ to evaluate the differences between human measurements and AI-based PlacentaVision measurements of the placental disc.

### PlacentaVision Study

PlacentaVision is a multi-site study to create AI-based models to assess placental morphology, features, and pathologies from digital photographs.^42^ We built a large dataset of digital placenta photos and clinical data on the placenta, mother, and offspring from singleton births at three large hospitals: Northwestern Memorial in Chicago, USA, UPMC Magee-Womens Hospital in Pittsburgh, USA, and Mbarara Regional Referral Hospital affiliated with the Mbarara University of Science and Technology (MUST) in Mbarara, Uganda. The Pennsylvania State University Institutional Review Board (IRB) served as the single IRB for all research in the US and gave ethical approval for this work (STUDY00020697); the MUST Research Ethics Committee reviewed and gave ethical approval for the research at MUST (HS3159ES).

In the current analysis, we use data from the gross examination conducted by trained individuals (pathology technicians at Northwestern; pathology technicians and research staff at Magee; research staff at MUST) and our PlacentaVision models. We developed a photography protocol (https://osf.io/9e6nm/overview) which was prospectively followed at Magee and MUST. Photographs from Northwestern followed the pathology department’s protocol, which generally aligned with PlacentaVision.

### Sample Size

Data from Northwestern were extracted from electronic medical records for all deliveries of singleton births with a placental pathology report from January 1, 2012 to December 31, 2022 (n=35,077). Data from Magee were from all participants in the Magee Obstetric Medical and Infant (MOMI) Biobank study (n=1371)^43,44^ with singleton births and placental pathology report. Data from MUST were all participants in the Placentas, Antibodies, and Child Outcomes (PACO) study (n=501) and all participants in our PlacentaVision study (n=1387). The total initial combined dataset was 38,336 births. We dropped 5011 records which were missing placental photographs (image file missing and/or error with photo including not an image of the full disc and cord), and an additional 666 for which the quality of the photograph or the ruler was too poor to see/read the ruler (e.g. glare in image; lines rubbed off ruler) and 92 which were missing a ruler.

Given our current goal of examining a single disc length and width, we removed those identified as fragmented/disrupted/not whole discs (n=1038) and those with multiple lobes (i.e., bilobed or accessory lobe(s); n=2618) because these have multiple measurements in the pathology exam. We also dropped those with velamentous or furcate umbilical cord insertion (n=555) because these photos show membranes spread out with the cord and PlacentaVision is not yet trained to segment membranes. Next, we dropped those with gestational age missing (n=209), <13 weeks (the lower limit of standard pathology exams, n=1), and >43 weeks (given the likelihood of error in gestational age, n=15). We then dropped those with missing length or width in the gross exam (n=119) and implausibly low (<5 cm, n=84) or implausibly high (>35 cm, n=80) length or width from gross exam or PlacentaVision data for a final analytic sample of 27,846.

### Gross placental examination

Northwestern Memorial Hospital is a tertiary care facility in Chicago, Il with approximately 12,000 births per year. The decision tree for sending placentas for pathological examination is provided in supplemental materials (Supplemental Figure 1) and yields ∼20% of placentas being sent. While clinical providers follow the decision tree, they also have the liberty to send placentas for any reason they feel is warranted. After delivery, all placentas are labeled in a waterproof container and placed in refrigeration near the labor and delivery ward. Placentas designated for pathology are picked up by the pathology department once a day and put in formalin (whole organ). Technicians and pathologist assistants complete the gross examination as it fits in their workflow. In cases when the placenta data is deemed urgent, exams can be conducted in <24 hours. In most cases, the exam occurs 24-72 hours after delivery. Technicians have a workstation in which they assess, measure, and sample the placental disc, membranes, and umbilical cord. Results are recorded in real time in the medical record software. Placentas not sent to pathology discarded from refrigeration after 3 days. The final pathology report, including the gross examination and histological diagnoses, was extracted for the PlacentaVision dataset.

UPMC Magee-Womens Hospital is a tertiary care facility in Pittsburgh, PA with approximately 10,000 births per year. Placentas included in the current analysis were collected as part of the Magee Obstetric Maternal & Infant (MOMI) database and biobank. Approximately 20% of women delivering at Magee-Womens Hospital enroll in the MOMI biobank study and provide written consent for data, placental photography, and specimen collection. Within 30 minutes after delivery, placentas from MOMI participants underwent research examination consisting of photographs, measurements, and tissue sampling. If a clinical pathology examination was ordered based on hospital guidelines (which include maternal, fetal, and placental indications) and/or provider judgement, it was then sent to the pathology department for full examination (approximately 50% of the MOMI/biobank cohort). The PlacentaVision Magee dataset included measurements of the placenta disc which were extracted from the pathology report. Since the Magee pathology department does not obtain photographs of every placenta, in contrast to Northwestern, the PlacentaVision dataset utilized images obtained by MOMI research staff.

At MUST, placentas are rarely examined by the pathology department and there are no perinatal pathologists (a situation which is similar in low-resource settings globally). For the current analysis, placenta data and images were used from all participants in the PACO and PlacentaVision studies. Placentas from enrolled research participants were collected at birth and taken to a designated research space where they were photographed and a gross exam was performed within 2 hours of birth. Trained technicians recorded gross exam information on paper, then transferred data to RedCap using designated study iPads. Samples of disc, cord, and membranes were collected and placed in labeled formalin containers for 24 hours. Samples were then trimmed, placed in tissue cassettes and transferred to the pathology department for processing. Histological diagnoses were conducted by perinatal pathologists in the US.

### Placenta photography

We developed a protocol for taking photographs of the maternal and fetal (with umbilical cord) sides of the placenta for PlacentaVision,^45^ adapted from the photographing procedure that was established at Northwestern (which started approximately 2010). In brief, each placenta should be blotted to remove excess blood and non-adherent clots then placed in the middle of a blue background (e.g. a blue cutting board). For the fetal side photograph, the membranes should be tucked underneath the disc and the cord should be laid off the disk at the point closest to the insertion point and wrapped around the edge without directly touching the disk. For the maternal side, the membranes and cord should be tucked underneath so that only the maternal surface is showing. For all photos, a ruler with centimeter markings should be placed on the blue board (not touching any part of disc or cord), which is later read by computer vision models to calculate placenta dimensions. Nothing else, except a study or patient identification number, should appear in the photograph.

All photographs were taken using digital cameras (not mobile devices or tablets). At **Northwestern**, the pathology department has fully furnished photography stations as part of the normal gross examination workflow. These stations include a professional pathology specimen photography system (Macropath, Milestone Medical, Kalamazoo, MI, USA) mounted on a stand that is positioned above the blue photography surface and placenta. Different rulers were used over the 10+ years; almost all were small white rulers. For many years of the project, a printed laminated paper ruler was used (as it was easy to replace). At **Magee**, photographs were taken specifically for the MOMI Biobank research project using a Canon model DS126741 camera. At the beginning of the study, research personnel were holding the camera and taking the photographs by hand. At some point, a camera stand was purchased and used to take photographs parallel to the surface and at a consistent distance from the placenta. This study used a large, black L-shaped ruler. For both the PACO and PlacentaVision studies at **MUST**, photographs were taken using a camera held by hand on a blue cutting board with a small white ruler. A Fujifilm FinePix XP130 digital camera was used for the PACO study and a Canon EOS Rebel T7 DSLR camera with an 18-55 mm lens was used for PlacentaVision.

Lighting varied between sites. Most notably, photographs in both Northwestern and Magee were taken in interior rooms with artificial light, while there was a large window in the space used for photography at MUST, and small portable lights were used as needed (e.g. when cloudy).

At Northwestern, placenta photographs are taken as part of the standard pathology exam and one photograph per side was available as part of the medical record. Similarly, at Magee a single photograph was taken for each side, but photographs were saved in the research data repository. Finally, at MUST, research assistants took multiple photographs for each side, and the highest quality image was selected by research personnel prior to analysis.

### Placental Morphology

Length and width, or axes of the placental disc, have been defined in various ways over time and often descriptions or directions in gross examination guidelines are somewhat vague. The Amsterdam consensus^31^ included the following definitions:

**Length**: “the maximal linear dimension”

**Width**: “the greatest dimension of the axis perpendicular to this [length] linear measurement”

**Gross examination**. Northwestern and Magee have gross exam protocols that describe how to measure length and width according to Amsterdam criteria. For the PACO and PlacentaVision studies at MUST, the protocol for research staff instructed measurement of “greatest diameter” and “other greatest diameter” with in-person training to take the second measurement perpendicular to the first.

**PlacentaVision Morphology Models.** PlacentaVision extracts measurements from placenta images through a two-stage process.^35,37^ First, it segments the placental disc, identifies the umbilical cord insertion point, and classifies the image as either the fetal or maternal side. Next, it computes key morphological features such as placental area, perimeter, long and short diameters, and cord insertion type. These steps are automated using deep learning models trained on annotated placenta images, combined with traditional computer vision algorithms, enabling consistent and objective analysis across diverse imaging conditions.

**Variables from PlacentaVision.** Morphology measurements performed by the PlacentaVision algorithm used the ruler in the image to calculate dimensions. The model calculated the **length** (cm) by identifying the greatest dimension between any two points on the disk perimeter and the **width** (cm) by identifying the greatest dimension at a perpendicular angle from the length, both following Amsterdam consensus definitions.^31^ The model measured several additional features and dimensions of the placenta not done (or feasible) in a human gross examination, including disk perimeter (cm), disk area (cm^2^), and a quantitative measure of elliptical disk shape (ratio). The model overlaid a fitted ellipse onto the disk for quantification of the elliptical shape of the placenta disk (Figure 1). Then, the areas inside and outside the ellipse (labeled as ‘1’ in the figure) were calculated and summed together. This sum is then divided by the area of the ellipse to obtain a shape ratio. We defined a shape ratio of >0.1 (10%) as irregular and ≤0.1 as regular.

**Figure 1.**
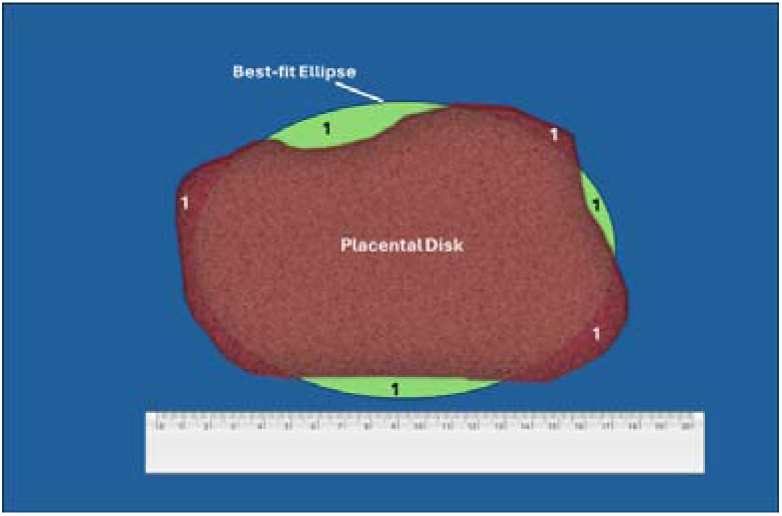
PlacentaVision algorithm calculation of shape ratio. To assess shape, the PlacentaVision algorithm calculates the best fit ellipse from the placental disk, then calculates the total amount of disk inside and outside the ellipse, labeled ‘1’ in the figure. This area is divided by the area of the ellipse to produce the shape ratio.

### Variables from pathology report and clinical data

Several variables were extracted from the pathology report for comparison with the PlacentaVision algorithm. For data from Northwestern and Magee, this involved extracting variables from the narrative text of the report. At MUST, data were entered in a RedCap form as individual variables. Numerical variables extracted from the pathology report included the disc dimensions in cm (disc length, width, and thickness) and placental weight (g). We also extracted disc shape (e.g. round, oval, irregular) from Northwestern and MUST; shape is not recorded at Magee in standard gross exams. Clinical and demographic variables were extracted from the medical record at Northwestern, obtained from the MOMI biobank data repository at Magee, and obtained from a combination of self-report and medical record extraction at MUST. These clinical variables included infant sex, gestational age at birth, infant birth weight, mode of delivery, maternal parity (MUST: only gravidity was available), maternal race and ethnicity (not recorded at MUST), and maternal age.

### Statistical analysis

We examined distributions for all continuous variables and frequencies for all categorical variables; summary statistics were calculated based on distribution type and report overall and by site. We used kernel density plots with a normal overlay to visualize continuous distributions and examined kurtosis and skewness to classify distributions as approximately Gaussian or not.

To examine the differences between human and PlacentaVision measurements, we calculated the difference in disc length, disc width, and cord insertion distance from the disc margin as: *PlacentaVision – gross exam = difference*. All values were in cm.

We then classified the magnitude of difference as similar (≥ −1 cm & ≤ 1cm); PlacentaVision longer (>1cm); or PlacentaVision shorter (<-1cm) and summarized the proportion in each category. We also calculated Lin’s Concordance Correlation Coefficient (CCC), which assesses agreement between variables by accounting for the deviation from a 45-degree line of agreement in addition to linear agreement. We further compared measurements using Bland-Altman analysis and plots. In this method, the difference in the 2 measurements was plotted against the mean of the 2 measurements, and limits of detection were calculated as 2 standard deviations from the mean difference.

For each set of analyses, we stratified by site (Northwestern, Magee, or MUST), shape (regular or irregular), infant sex (male or female), and gestational age at birth (preterm [<37 weeks] or term [≥37 weeks]) and tested mean differences with linear regression. Finally, we examined potential reasons for the discrepancies between the methods by reviewing edge cases where measurements were >|10cm| different for length and width.

## Results

### Participant characteristics

The total sample of participants included in this analysis had mean maternal age of 31.6 years, a median parity of 1, a median gestational age at birth of 39 weeks, and 36% delivered by Cesarean (Table 1). Some characteristics varied by site. The mean maternal age was younger at MUST compared to the other sites. Inherently due to population, all of the participants at MUST were Black compared to 14.5% at Northwestern and 15.4% at Magee. At Magee, 78.1% of participants were of White race, while Northwestern participants were the most racially and ethnically diverse. Primiparous births were much higher at Northwestern compared to the other sites. At Northwestern, 23.9% of placentas were from preterm birth (<37 weeks), while these numbers were lower at Magee (15.0%) and MUST (12.5%). Similarly, prevalence of low birth weight (<2500 g) was higher at Northwestern (20.3%) compared with Magee (13.6%) and MUST (8.1%).

**Table 1.** Maternal, infant, and pregnancy characteristics by hospital.

| Characteristics | Total Sample<br>(N=27,846) | Hospital |  |  |
| --- | --- | --- | --- | --- |
|  |  | Northwestern<br>(N=24,933) | Magee<br>(N=1198) | MUST<br>(N=1715) |
| | | Mean ± SD or N (%) | | |
| Maternal age (years) | 31.6 (5.6) | 32.0 (5.4) | 30.6 (5.3) | 26.6 (6.0) |
| Maternal race |  |  |  |  |
| White | 14,296 (51.3) | 13,361 (53.6) | 935 (78.1) | 0 |
| Black | 5,525 (19.8) | 3,625 (14.5) | 185 (15.4) | 1,715 (100) |
| Asian | 2,228 (8.0) | 2,178 (8.7) | 50 (4.2) | 0 |
| Other<sup>1</sup> | 5,796 (20.8) | 5,769 (23.1) | 28 (2.3) | 0 |
| Maternal ethnicity |  |  |  |  |
| Hispanic | 4,712 (16.9) | 4,693 (18.8) | 19 (1.6) | 0 |
| Non-Hispanic<sup>2</sup> | 23,134 (83.1) | 20,240 (81.2) | 1,179 (98.4) | 1,715 (100) |
| Parity<sup>3</sup> (median, IQR) | 1 (1, 2) | 1 (1, 2) | 2 (2, 3) | 2 (1, 4) |
| Primiparous birth | 15,032 (57.7) | 14,436 (62.2) | 21 (1.9) | 575 (33.6) |
| Gestational age (wk)<br>(median, IQR) | 39.0 (37.0, 40.0) | 39.0 (37.0, 40.0) | 39.0 (37.4, 39.6) | 39.0 (38.0, 40.0) |
| Preterm birth | 6,349 (22.8) | 5,955 (23.9) | 180 (15.0) | 214 (12.5) |
| Cesarean delivery<br>(Birth mode) | 9,815 (36.1) | 8,708 (35.8) | 472 (39.5) | 635 (37.1) |
| Infant sex (male) | 14,130 (53.1) | 12,595 (53.2) | 620 (52.0) | 915 (53.5) |
| Infant birth weight (g) | 3036 (737) | 3025 (753) | 3156 (653) | 3115 (515) |
| Low birth weight<br>(<2500 g) | 5,319 (19.3) | 5,018 (20.3) | 163 (13.6) | 138 (8.1) |
| Placenta weight (g) | 432 (115) | 433 (117) | 397 (99) | 449 (99) |

<sup>1</sup> Includes Other, American Indian, Alaskan Native, Native Hawaiian, Other Pacific Islander, Declined, and Unknown responses
<sup>2</sup> Includes Declined, Unspecified, Unknown, and missing responses
<sup>3</sup> Gravidity used for MUST studies because parity (before current birth) unavailable

### Morphology measured by gross examination and PlacentaVision

The mean differences between length and width measured in the gross exam compared with length and width calculated by the PlacentaVision algorithm were small (Table 2). PlacentaVision length was 0.57 cm greater (SD 2.19) and width was 0.25 cm greater (SD 1.85) compared with the gross exam. The distributions were largely overlapping, but notably the gross exam measurement distributions show digit preference for rounded numbers (Supplemental Figure 2). The mean difference was the highest at Northwestern, followed by MUST and Magee. The mean difference in measurements at Magee was 0 with a similar standard deviation (i.e., a similar number of discrepancies where PlacentaVision was longer or shorter than the gross exam). Mean differences between gross exam and PlacentaVision in both length and width were higher in placentas with an irregular shape (1.53 and 0.42 cm, respectively) compared to a regular shape (0.45 and 0.23 cm, respectively). Results for male and female placentas were similar. Preterm had slightly larger differences in the measurements compared to term placentas.

**Table 2.**
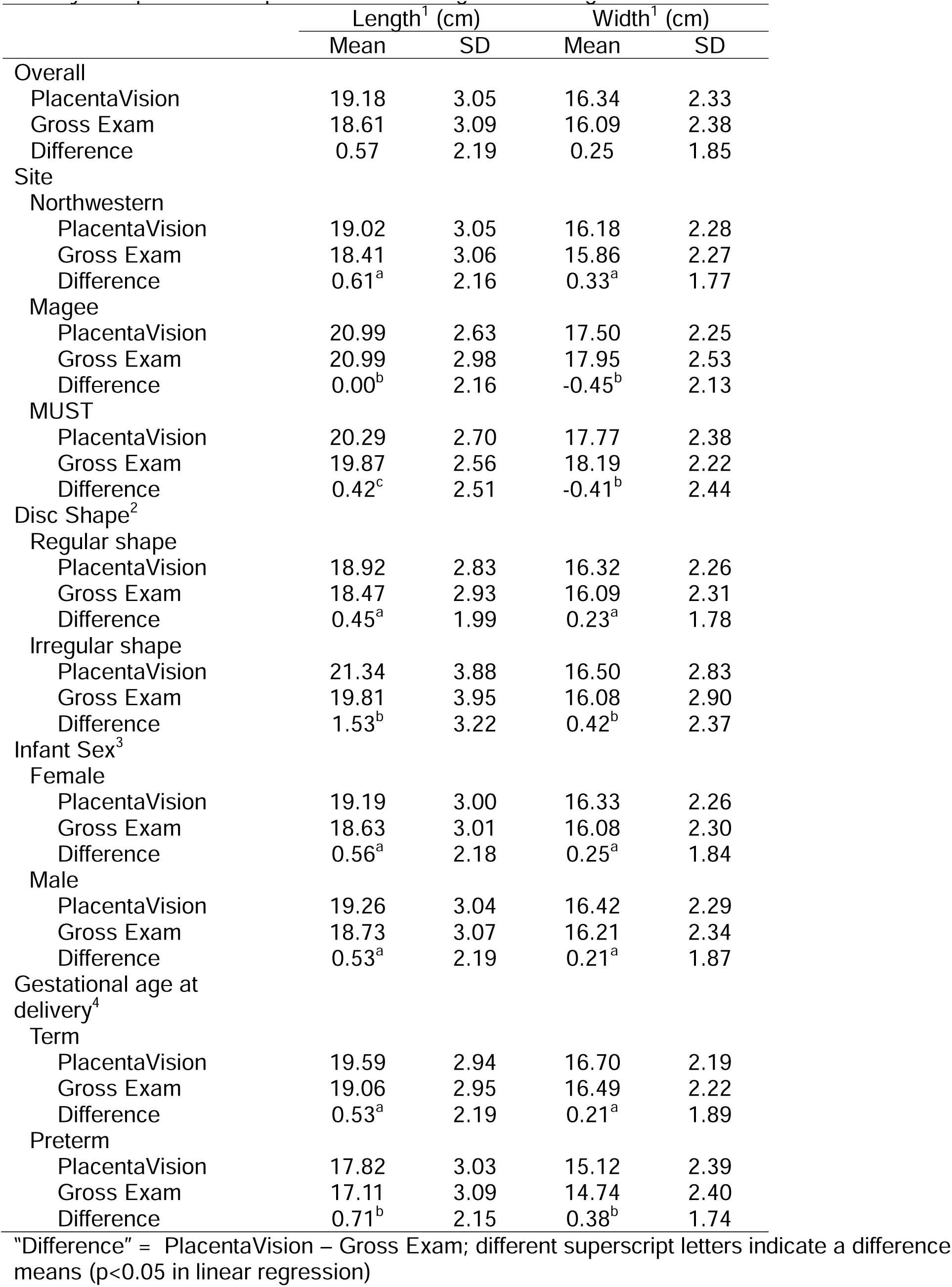
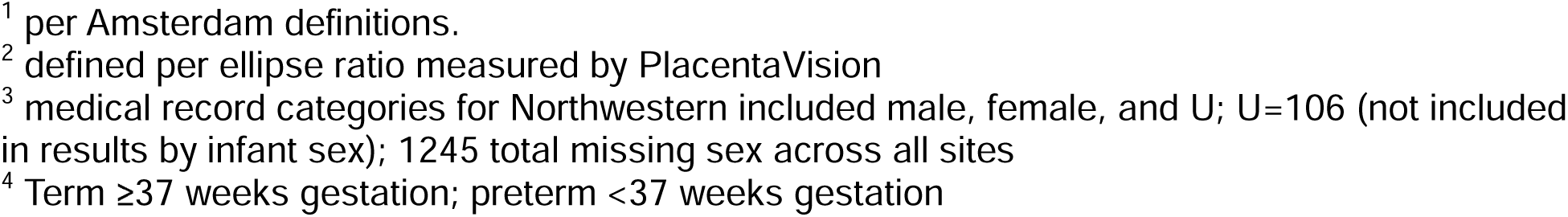
Placental morphology from PlacentaVision and gross pathology examination, overall and by site, placenta shape, infant sex, and gestational age, n=27,846.

|  | Length<sup>1</sup> (cm) |  | Width<sup>1</sup> (cm) |  |
| --- | --- | --- | --- | --- |
|  | Mean | SD | Mean | SD |
| Overall |  |  |  |  |
| PlacentaVision | 19.18 | 3.05 | 16.34 | 2.33 |
| Gross Exam | 18.61 | 3.09 | 16.09 | 2.38 |
| Difference | 0.57 | 2.19 | 0.25 | 1.85 |
| Site |  |  |  |  |
| Northwestern |  |  |  |  |
| PlacentaVision | 19.02 | 3.05 | 16.18 | 2.28 |
| Gross Exam | 18.41 | 3.06 | 15.86 | 2.27 |
| Difference | 0.61<sup>a</sup> | 2.16 | 0.33<sup>a</sup> | 1.77 |
| Magee |  |  |  |  |
| PlacentaVision | 20.99 | 2.63 | 17.50 | 2.25 |
| Gross Exam | 20.99 | 2.98 | 17.95 | 2.53 |
| Difference | 0.00<sup>b</sup> | 2.16 | -0.45<sup>b</sup> | 2.13 |
| MUST |  |  |  |  |
| PlacentaVision | 20.29 | 2.70 | 17.77 | 2.38 |
| Gross Exam | 19.87 | 2.56 | 18.19 | 2.22 |
| Difference | 0.42<sup>c</sup> | 2.51 | -0.41<sup>b</sup> | 2.44 |
| Disc Shape<sup>2</sup> |  |  |  |  |
| Regular shape |  |  |  |  |
| PlacentaVision | 18.92 | 2.83 | 16.32 | 2.26 |
| Gross Exam | 18.47 | 2.93 | 16.09 | 2.31 |
| Difference | 0.45<sup>a</sup> | 1.99 | 0.23<sup>a</sup> | 1.78 |
| Irregular shape |  |  |  |  |
| PlacentaVision | 21.34 | 3.88 | 16.50 | 2.83 |
| Gross Exam | 19.81 | 3.95 | 16.08 | 2.90 |
| Difference | 1.53<sup>b</sup> | 3.22 | 0.42<sup>b</sup> | 2.37 |
| Infant Sex<sup>3</sup> |  |  |  |  |
| Female |  |  |  |  |
| PlacentaVision | 19.19 | 3.00 | 16.33 | 2.26 |
| Gross Exam | 18.63 | 3.01 | 16.08 | 2.30 |
| Difference | 0.56<sup>a</sup> | 2.18 | 0.25<sup>a</sup> | 1.84 |
| Male |  |  |  |  |
| PlacentaVision | 19.26 | 3.04 | 16.42 | 2.29 |
| Gross Exam | 18.73 | 3.07 | 16.21 | 2.34 |
| Difference | 0.53<sup>a</sup> | 2.19 | 0.21<sup>a</sup> | 1.87 |
| Gestational age at delivery<sup>4</sup> |  |  |  |  |
| Term |  |  |  |  |
| PlacentaVision | 19.59 | 2.94 | 16.70 | 2.19 |
| Gross Exam | 19.06 | 2.95 | 16.49 | 2.22 |
| Difference | 0.53<sup>a</sup> | 2.19 | 0.21<sup>a</sup> | 1.89 |
| Preterm |  |  |  |  |
| PlacentaVision | 17.82 | 3.03 | 15.12 | 2.39 |
| Gross Exam | 17.11 | 3.09 | 14.74 | 2.40 |
| Difference | 0.71<sup>b</sup> | 2.15 | 0.38<sup>b</sup> | 1.74 |

“Difference” = PlacentaVision – Gross Exam; different superscript letters indicate a difference means ( $p < 0.05$ in linear regression)
<sup>1</sup> per Amsterdam definitions.
<sup>2</sup> defined per ellipse ratio measured by PlacentaVision
<sup>3</sup> medical record categories for Northwestern included male, female, and U; U=106 (not included in results by infant sex); 1245 total missing sex across all sites
<sup>4</sup> Term $\geq 37$ weeks gestation; preterm $< 37$ weeks gestation

The examination of the proportion of placentas with close alignment of measurements (within 1 cm) or longer or shorter measurements (>|1| cm) showed a similar pattern (Table 3). Overall, 45% of length measurements and 50% of width measurements were within 1 cm of each other for PlacentaVision and the gross exam. Length and width both had a higher proportion of measurements in which PlacentaVision was longer compared to shorter than the gross exam. Northwestern and MUST had a similar pattern to the overall results, while Magee had fewer placentas with close measurement alignment and an approximately even proportion of placentas with PlacentaVision being longer or shorter than the gross exam. For irregular compared to regular shapes, there was a higher proportion of measurements in which PlacentaVision was longer than the gross exam. Comparison by infant sex and gestational age at birth found overall similarities in these categories.

**Table 3.**
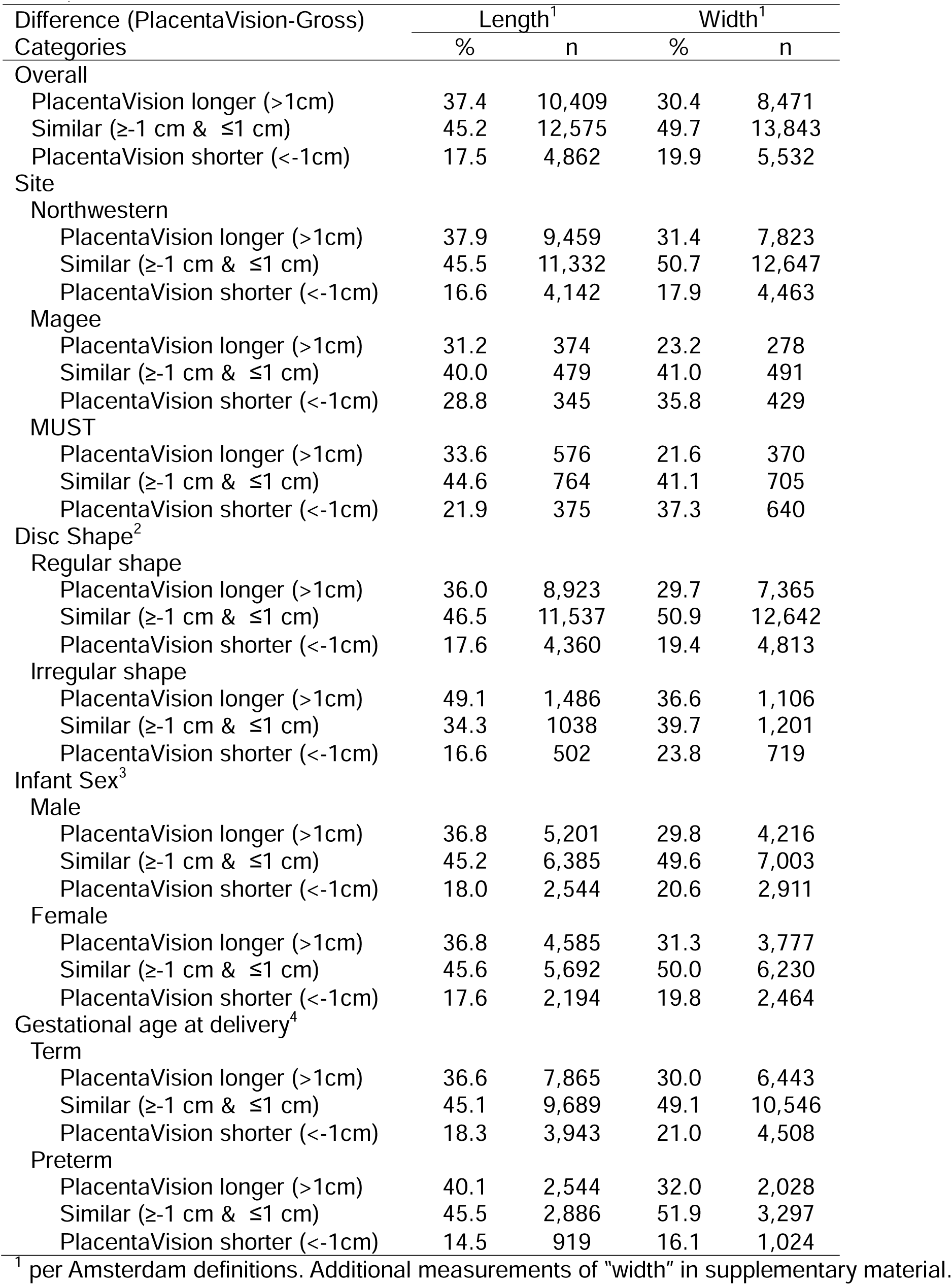
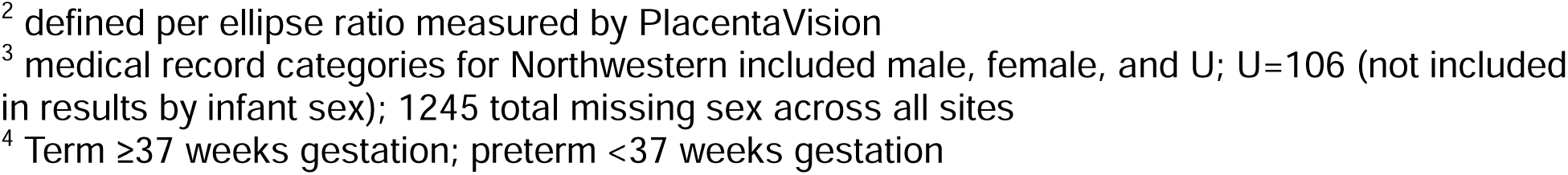
Proportional differences in placental morphology from the gross pathology examination vs PlacentaVision, overall and by placenta shape, site, infant sex, and gestational age, n=27,846.

| Difference (PlacentaVision-Gross)<br>Categories | Length<sup>1</sup> |  | Width<sup>1</sup> |  |
| --- | --- | --- | --- | --- |
|  | % | n | % | n |
| Overall |  |  |  |  |
| PlacentaVision longer (>1cm) | 37.4 | 10,409 | 30.4 | 8,471 |
| Similar (≥-1 cm & ≤1 cm) | 45.2 | 12,575 | 49.7 | 13,843 |
| PlacentaVision shorter (<-1cm) | 17.5 | 4,862 | 19.9 | 5,532 |
| Site |  |  |  |  |
| Northwestern |  |  |  |  |
| PlacentaVision longer (>1cm) | 37.9 | 9,459 | 31.4 | 7,823 |
| Similar (≥-1 cm & ≤1 cm) | 45.5 | 11,332 | 50.7 | 12,647 |
| PlacentaVision shorter (<-1cm) | 16.6 | 4,142 | 17.9 | 4,463 |
| Magee |  |  |  |  |
| PlacentaVision longer (>1cm) | 31.2 | 374 | 23.2 | 278 |
| Similar (≥-1 cm & ≤1 cm) | 40.0 | 479 | 41.0 | 491 |
| PlacentaVision shorter (<-1cm) | 28.8 | 345 | 35.8 | 429 |
| MUST |  |  |  |  |
| PlacentaVision longer (>1cm) | 33.6 | 576 | 21.6 | 370 |
| Similar (≥-1 cm & ≤1 cm) | 44.6 | 764 | 41.1 | 705 |
| PlacentaVision shorter (<-1cm) | 21.9 | 375 | 37.3 | 640 |
| Disc Shape<sup>2</sup> |  |  |  |  |
| Regular shape |  |  |  |  |
| PlacentaVision longer (>1cm) | 36.0 | 8,923 | 29.7 | 7,365 |
| Similar (≥-1 cm & ≤1 cm) | 46.5 | 11,537 | 50.9 | 12,642 |
| PlacentaVision shorter (<-1cm) | 17.6 | 4,360 | 19.4 | 4,813 |
| Irregular shape |  |  |  |  |
| PlacentaVision longer (>1cm) | 49.1 | 1,486 | 36.6 | 1,106 |
| Similar (≥-1 cm & ≤1 cm) | 34.3 | 1038 | 39.7 | 1,201 |
| PlacentaVision shorter (<-1cm) | 16.6 | 502 | 23.8 | 719 |
| Infant Sex<sup>3</sup> |  |  |  |  |
| Male |  |  |  |  |
| PlacentaVision longer (>1cm) | 36.8 | 5,201 | 29.8 | 4,216 |
| Similar (≥-1 cm & ≤1 cm) | 45.2 | 6,385 | 49.6 | 7,003 |
| PlacentaVision shorter (<-1cm) | 18.0 | 2,544 | 20.6 | 2,911 |
| Female |  |  |  |  |
| PlacentaVision longer (>1cm) | 36.8 | 4,585 | 31.3 | 3,777 |
| Similar (≥-1 cm & ≤1 cm) | 45.6 | 5,692 | 50.0 | 6,230 |
| PlacentaVision shorter (<-1cm) | 17.6 | 2,194 | 19.8 | 2,464 |
| Gestational age at delivery<sup>4</sup> |  |  |  |  |
| Term |  |  |  |  |
| PlacentaVision longer (>1cm) | 36.6 | 7,865 | 30.0 | 6,443 |
| Similar (≥-1 cm & ≤1 cm) | 45.1 | 9,689 | 49.1 | 10,546 |
| PlacentaVision shorter (<-1cm) | 18.3 | 3,943 | 21.0 | 4,508 |
| Preterm |  |  |  |  |
| PlacentaVision longer (>1cm) | 40.1 | 2,544 | 32.0 | 2,028 |
| Similar (≥-1 cm & ≤1 cm) | 45.5 | 2,886 | 51.9 | 3,297 |
| PlacentaVision shorter (<-1cm) | 14.5 | 919 | 16.1 | 1,024 |

<sup>1</sup> per Amsterdam definitions. Additional measurements of "width" in supplementary material.
<sup>2</sup> defined per ellipse ratio measured by PlacentaVision
<sup>3</sup> medical record categories for Northwestern included male, female, and U; U=106 (not included in results by infant sex); 1245 total missing sex across all sites
<sup>4</sup> Term $\geq 37$ weeks gestation; preterm $< 37$ weeks gestation

**Table 4.**
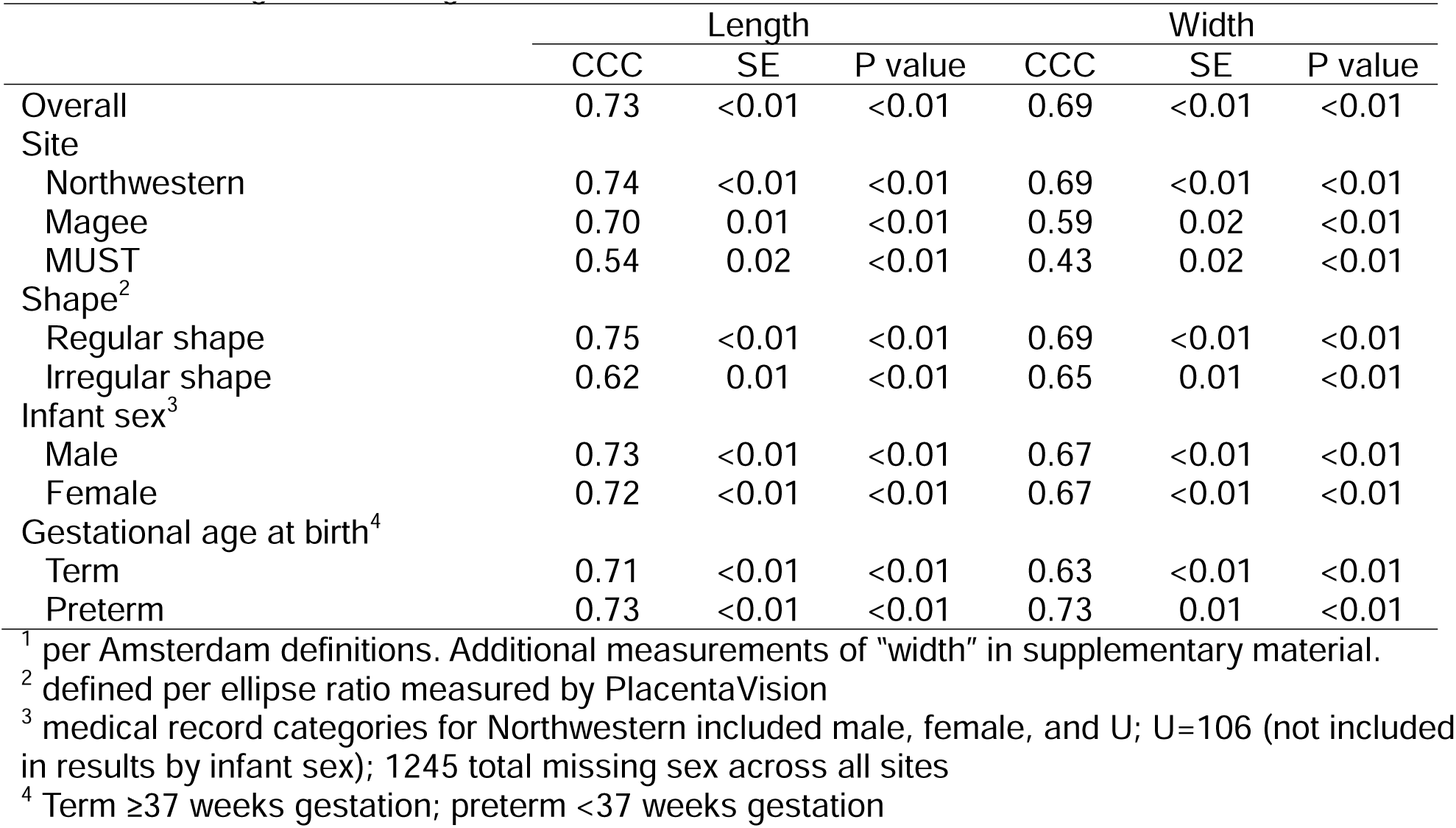
Lin’s concordance correlation coefficients (CCC) for placental morphology measured from PlacentaVision and the gross pathology examination, overall and by placenta shape, site, infant sex, and gestational age, n=27,846

|  | Length |  |  | Width |  |  |
| --- | --- | --- | --- | --- | --- | --- |
|  | CCC | SE | P value | CCC | SE | P value |
| Overall | 0.73 | <0.01 | <0.01 | 0.69 | <0.01 | <0.01 |
| Site |  |  |  |  |  |  |
| Northwestern | 0.74 | <0.01 | <0.01 | 0.69 | <0.01 | <0.01 |
| Magee | 0.70 | 0.01 | <0.01 | 0.59 | 0.02 | <0.01 |
| MUST | 0.54 | 0.02 | <0.01 | 0.43 | 0.02 | <0.01 |
| Shape<sup>2</sup> |  |  |  |  |  |  |
| Regular shape | 0.75 | <0.01 | <0.01 | 0.69 | <0.01 | <0.01 |
| Irregular shape | 0.62 | 0.01 | <0.01 | 0.65 | 0.01 | <0.01 |
| Infant sex<sup>3</sup> |  |  |  |  |  |  |
| Male | 0.73 | <0.01 | <0.01 | 0.67 | <0.01 | <0.01 |
| Female | 0.72 | <0.01 | <0.01 | 0.67 | <0.01 | <0.01 |
| Gestational age at birth<sup>4</sup> |  |  |  |  |  |  |
| Term | 0.71 | <0.01 | <0.01 | 0.63 | <0.01 | <0.01 |
| Preterm | 0.73 | <0.01 | <0.01 | 0.73 | 0.01 | <0.01 |

<sup>1</sup> per Amsterdam definitions. Additional measurements of "width" in supplementary material.
<sup>2</sup> defined per ellipse ratio measured by PlacentaVision
<sup>3</sup> medical record categories for Northwestern included male, female, and U; U=106 (not included in results by infant sex); 1245 total missing sex across all sites
<sup>4</sup> Term ≥37 weeks gestation; preterm <37 weeks gestation

Lin’s concordance analysis found an overall correlation of 0.73 for length and 0.69 for width (p<0.01) and scatterplots show a linear relationship (Supplemental Figure 3). Northwestern and Magee had similar correlation values while the correlations for MUST were much lower (<0.55). Irregular shape had lower correlation values compared to regular shape and results were similar by infant sex and gestational age at birth (all between 0.63 to 0.73). Bland Altman analysis (Figure 2) revealed the limits of agreement range was wider for length vs. width differences (−3.7 to 4.9 and −3.4 to 3.9 cm, respectively). The error in length and width measurements was evenly distributed across placenta size and across positive and negative differences, without indication of bias. Results by site are shown in Supplemental Figure 4. After manual review of cases with differences >|10|cm, we found PlacentaVision was longer or short due to ruler reading errors or longer due membranes in the photo (appearing as part of the disc) (Supplemental Figure 5). We also identified clear errors in the gross exam that appear to be data entry errors (e.g. where you can see from the ruler that the disc was approximately 20 cm but 30 cm was recorded). Finally, Supplemental Table 1 includes descriptive results of mean area and perimeter from PlacentaVision, values that are not measured or calculated in a gross exam.

**Figure 2.**
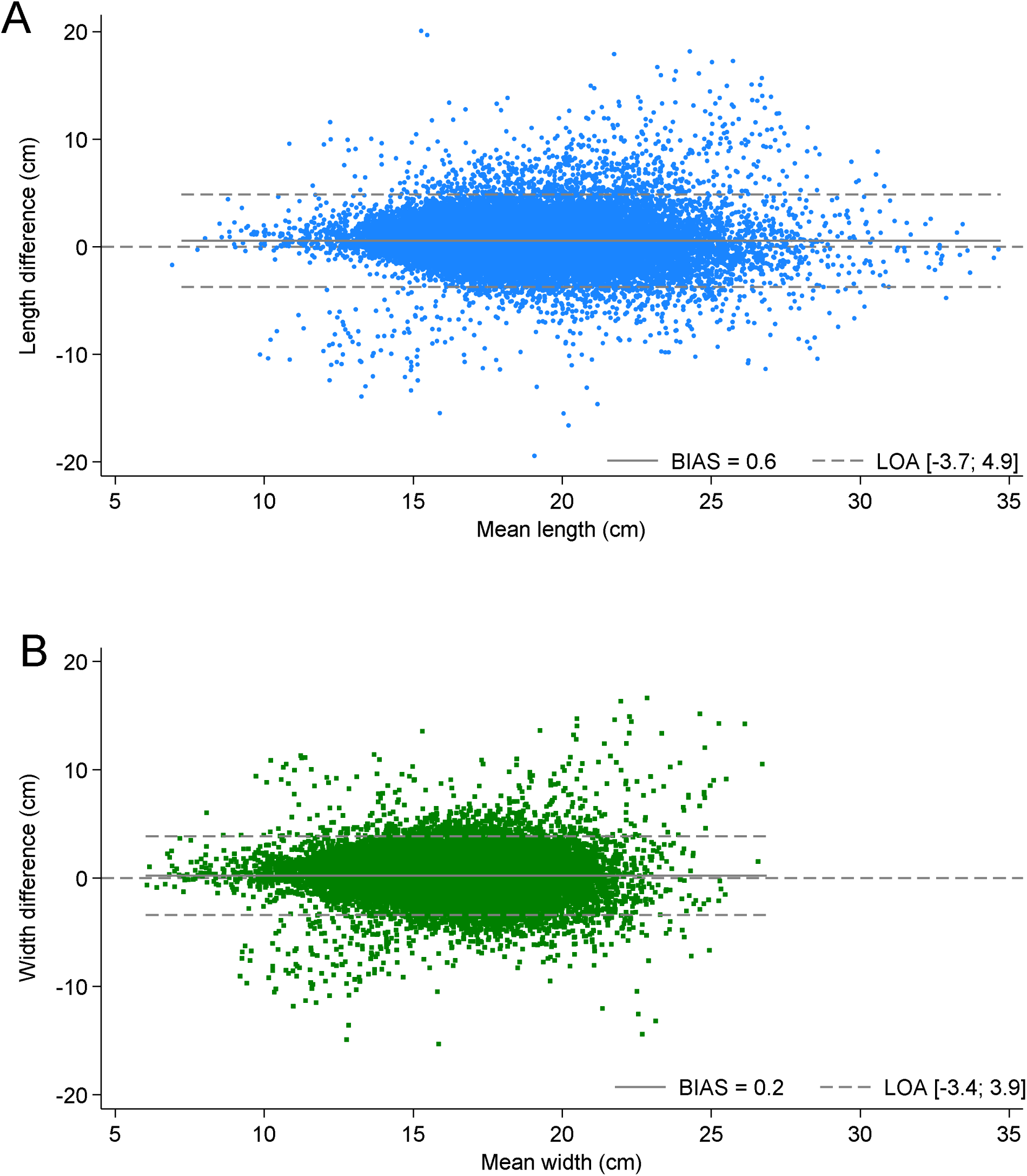
Bland Altman Plots of PlacentaVision vs. gross pathology examination measurement of (A) disc length and (B) disc width (n=27,846). LOA=Limits of agreement.

## Discussion

In this large study of placentas at 2 hospitals in the US and 1 in Uganda, AI-based PlacentaVision measurements were less than a cm from human measurements of disc length and width and approximately half of all measurements were within 1 cm of each other. Error was different by hospital with Northwestern having the largest mean difference between PlacentaVision and human measurements. Discs with irregular shape resulted in a much larger difference between PlacentaVision and human measurements compared to those with a round or oval shape. Differences were slightly larger for preterm vs. term placentas and there was no difference by sex. Correlation between the PlacentaVision and human measurements was high for length and width overall (0.7) and by categories, but MUST had the lowest correlations (length 0.5; width 0.4). Bland Altman analysis found 95% limits of agreement to be within 5 cm for length and within 4 cm for width, with a relatively even pattern of error across placenta size.

While the mean difference between PlacentaVision and human length and width measurements was small, the range of the difference was approximately −20 to 20 cm. In examining reasons for the error, we did not have data to document why a human measurement was wrong. In most cases with differences >10 cm and when PlacentaVision was correct, it was apparent that numbers were improperly recorded. For example, with a human length measurement of 32 cm and a (correct) PlacentaVision measurement of 23 cm we inferred that the numbers were inadvertently flipped during data entry. There were many cases, when PlacentaVision and human measurements were different by several cm with no obvious cause. However, we speculate that for irregularly shaped placentas, it is challenging for a person to identify the “true” length and width, and this led to the greater difference we observed between PlacentaVision and humans for irregular vs. regular discs.

Others have similarly acknowledged that “when the placenta is irregularly shaped, there may be substantial inter-observer variability in choosing a major and minor axis to measure.^46^ Furthermore, in the *Pathology of the Placenta*, authors recommended “a simple surface photograph, from which precise measurements can be made, and measurement methods improve as technology advances.”^34^ Our PlacentaVision project aims to make this recommendation a reality.

PlacentaVision models also had errors for two key reasons – the reading of the ruler was incorrect (such as not finding the markings) or there were membranes in the photo which appeared to the model as the disc (and were measured as such). The models we created for PlacentaVision are primarily limited by the mismatch between the variety of images in the training data and those encountered during real-world application. The models were trained and evaluated on a subset of data from the sites, and the application included images of widely varying quality (particularly from Northwestern, and most of the ruler errors were on images at this site). Despite the use of robust training and evaluation methods, a performance gap remains.

Furthermore, although the modular design and the integration of deep learning with traditional computer vision algorithms improve interpretability to some extent, the feature identification models in PlacentaVision—like most deep learning approaches—still lack detailed explainability. As a result, diagnosing how and why the model fails in certain cases remains challenging. Computer models can inherently create a more consistent definition to remove variability in characterization of shape and measurement of axes.

Mean length (19.18 cm) and width (16.34 cm) of placentas in our study are similar to those reported in the literature. Placentas from Saudi Arabia tend to have similar to slightly smaller dimensions to those measured in our study^7,8^, while placentas from Finland tended to be slightly larger in both length and width^12,16,18–21,23,26,27,47,48^. While dimensions were similar, mean placental weight in our study was smaller than weights reported in literature describing placental dimensions in generally healthy populations in Helsinki, Finland^12,16,18–21,23,26,27,47,48^ and slightly smaller than generally healthy placentas described from Saudi Arabia^7,8^. Many of these studies do not report how the placenta was weighed and therefore these higher values may be untrimmed placentas. The placentas measured in our study represent a wide variety of clinical outcomes, but the largest proportion were those sent to pathology because of some adverse outcome (which includes smaller preterm placentas and large but thin and irregular placentas). This may explain the similarity in dimensions but difference in weights measured in our study with those reported in generally healthy populations. Many of these studies were retrospective studies using data extracted from hospital records; length was typically defined as the “maximal diameter” of the placental surface and the “lesser diameter bisecting it at right angles” was considered the breadth. The prospective studies gave similar descriptions but noted that the placentas were measured with the cotyledons facing up, and in one case described that a transparent ruler was use used to measure the length after the longest diameter was determined, “by eye”.^12^ Additionally, many of the retrospective cohort studies from Barker and colleagues used hospital records where placental data were routinely collected in Finland, Holland and the UK, from 1924-1944. As such, baseline health indicators and particularly body mass index have certainly changed, underscoring the need for updated research.

Strengths of our study include the large sample size and multiple sites. As well, we compared the two methods of morphology measurements in multiple ways. The main limitation of the study is that the placement of the ruler for measurement was not documented as part the human gross exams, therefore when values misaligned and PlacentaVision was correct, it was difficult to know why the human measurement was wrong.

## Conclusion

In our study, PlacentaVision measurements from digital photos were extremely close to human measurements of the length and width of the placental disc, standard morphology measurements in gross exams which are important due to their link to health outcomes. Further, PlacentaVision can quantify additional useful parameters such as area and shape. Our results suggest that human error differs by site (presumably due to training and experience), and that people have difficulty identifying the length and width for irregular shapes. As well, there is error in data entry in electronic medical records. PlacentaVision models had errors due to incorrect ruler reading or membranes in the photo, and we are working to update AI models to correct these issues. Future studies should photograph human measurement for direct comparison of where rulers are placed on the disc, and qualitative research should be conducted to assess reasons for error and acceptability of using AI-based tools for measurements. PlacentaVision can automate, standardize, and expand morphological characterization of the human placenta and has the potential to improve gross examination and facilitate large-scale data collection.

## Data Availability

Data produced in the present study were shared from sites with investigators at Penn State via legal data use agreements and are not available publicly.

## Acknowledgements

We express our appreciation to the many members of the research team at each site who collected data, took photographs, and managed complicated datasets including Arnold Kamugisha (MUST), Immaculate Ninsiima (MUST), Jeannette Wellman (Magee-Womens Research Institute), and Aisha Owens (Northwestern). We also thank Drucilla J. Roberts, Professor Emerita of Pathology at Harvard Medical School, for her contributions to the PlacentaVision project and for conducting the majority of the pathology reports for placentas collected at MUST.

---

**Figure S1.**
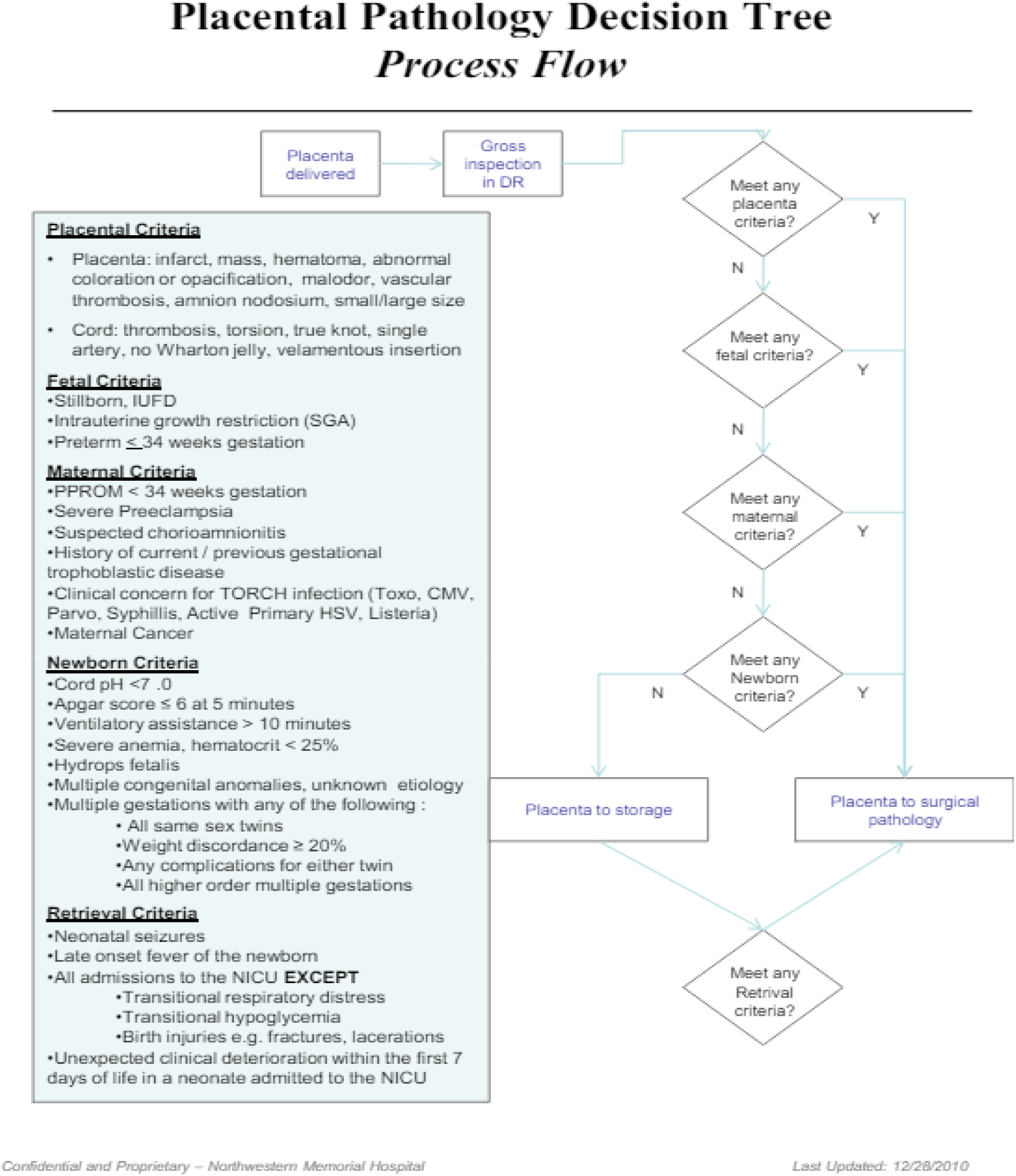
Placental pathology decision tree from Northwestern Memorial Hospital Abbreviations: APGAR= appearance, pulse, grimace, activity, and respiration; CMV=cytomegalovirus; DR=delivery room; HSV=herpes simplex virus; IUFD=intrauterine fetal demise; N=no; NICU=neonatal intensive care unit; PPROM=preterm premature rupture of membranes; SGA=small for gestational age; Y=yes

**Figure S2.**
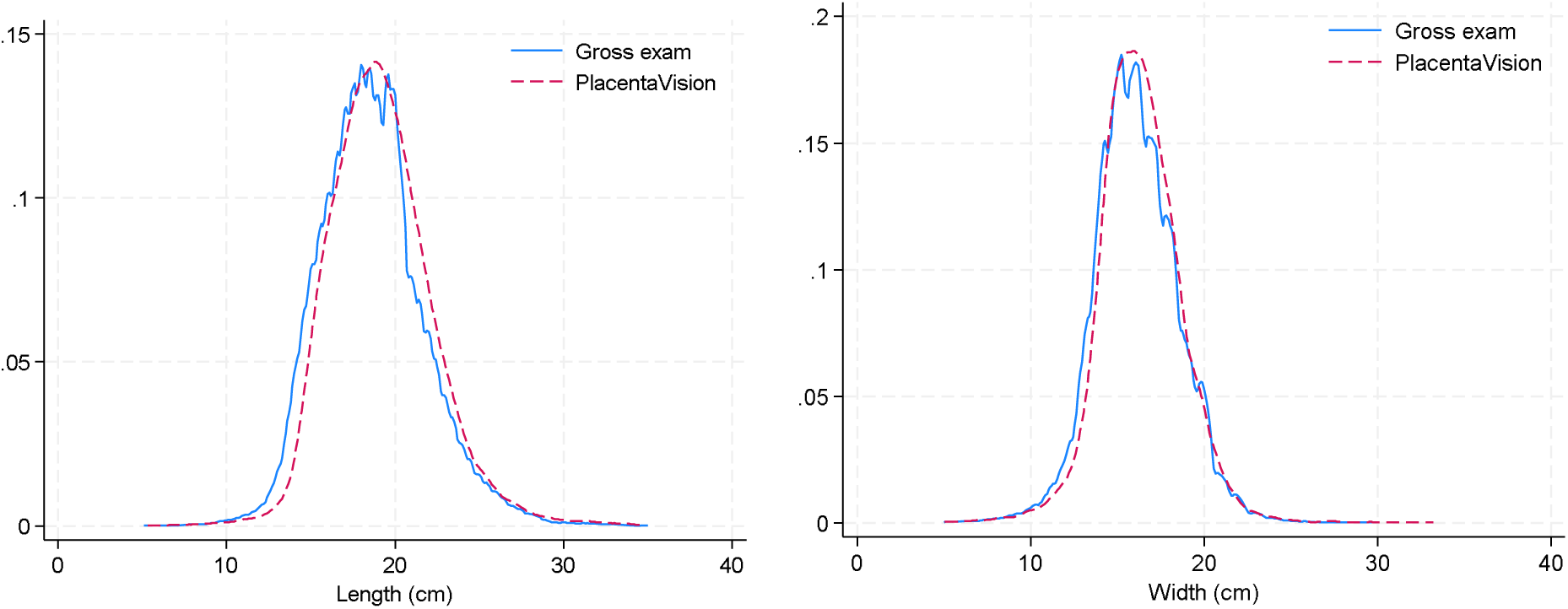
Kernel density plots of gross exam and PlacentaVision (by Amsterdam definition) length and width

**Figure S3.**
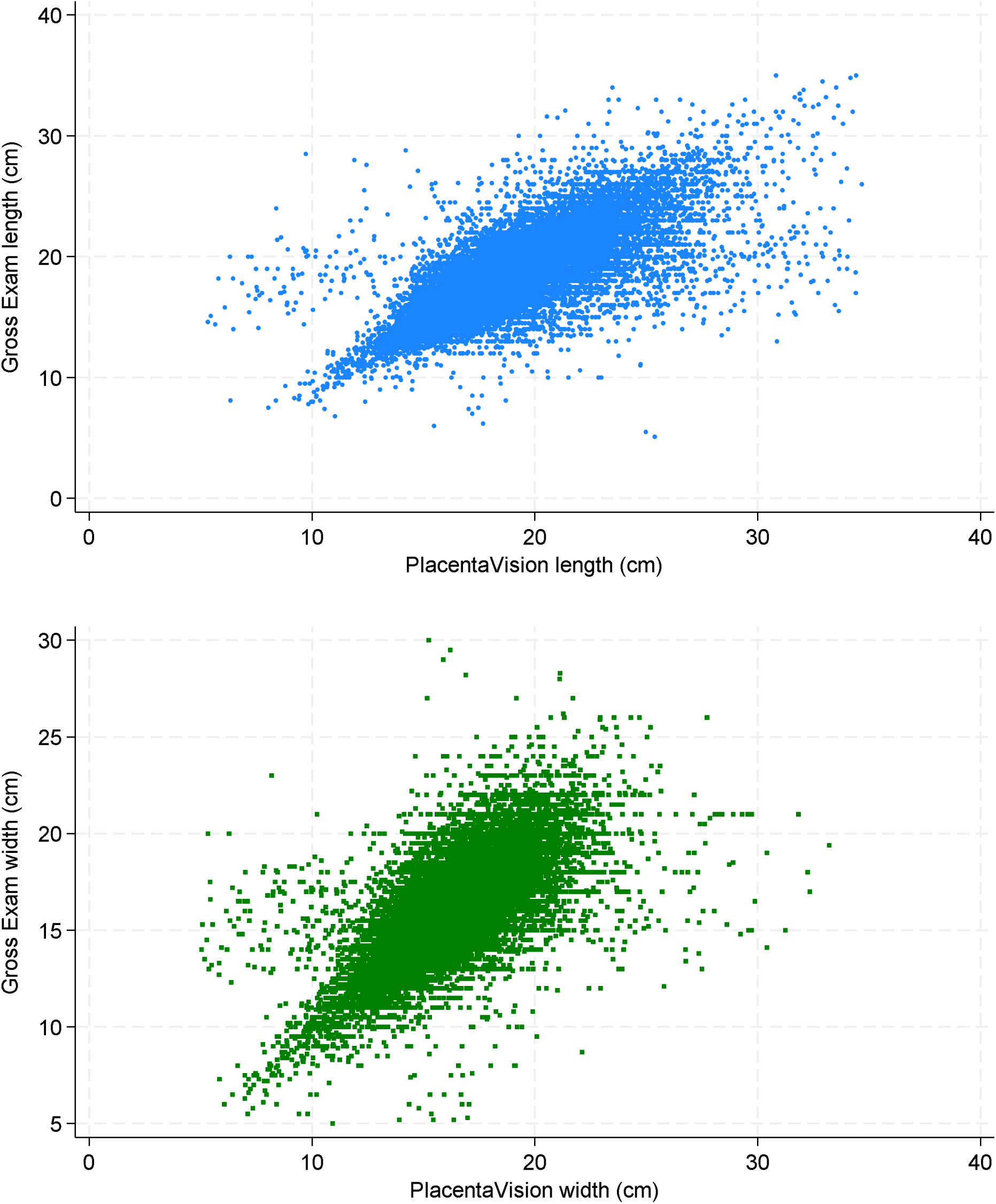
Scatter plots of gross exam and PlacentaVision (by Amsterdam definition) length and width

**Figure S4.**
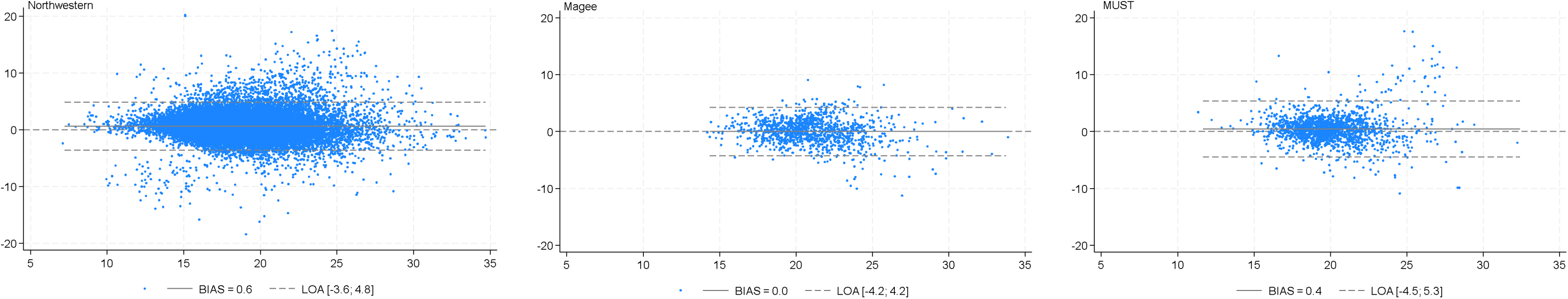
Bland Altman Plots of PlacentaVision vs. gross pathology examination measurement of disc length by site. LOA=Limits of agreement.

**Figure S5.**
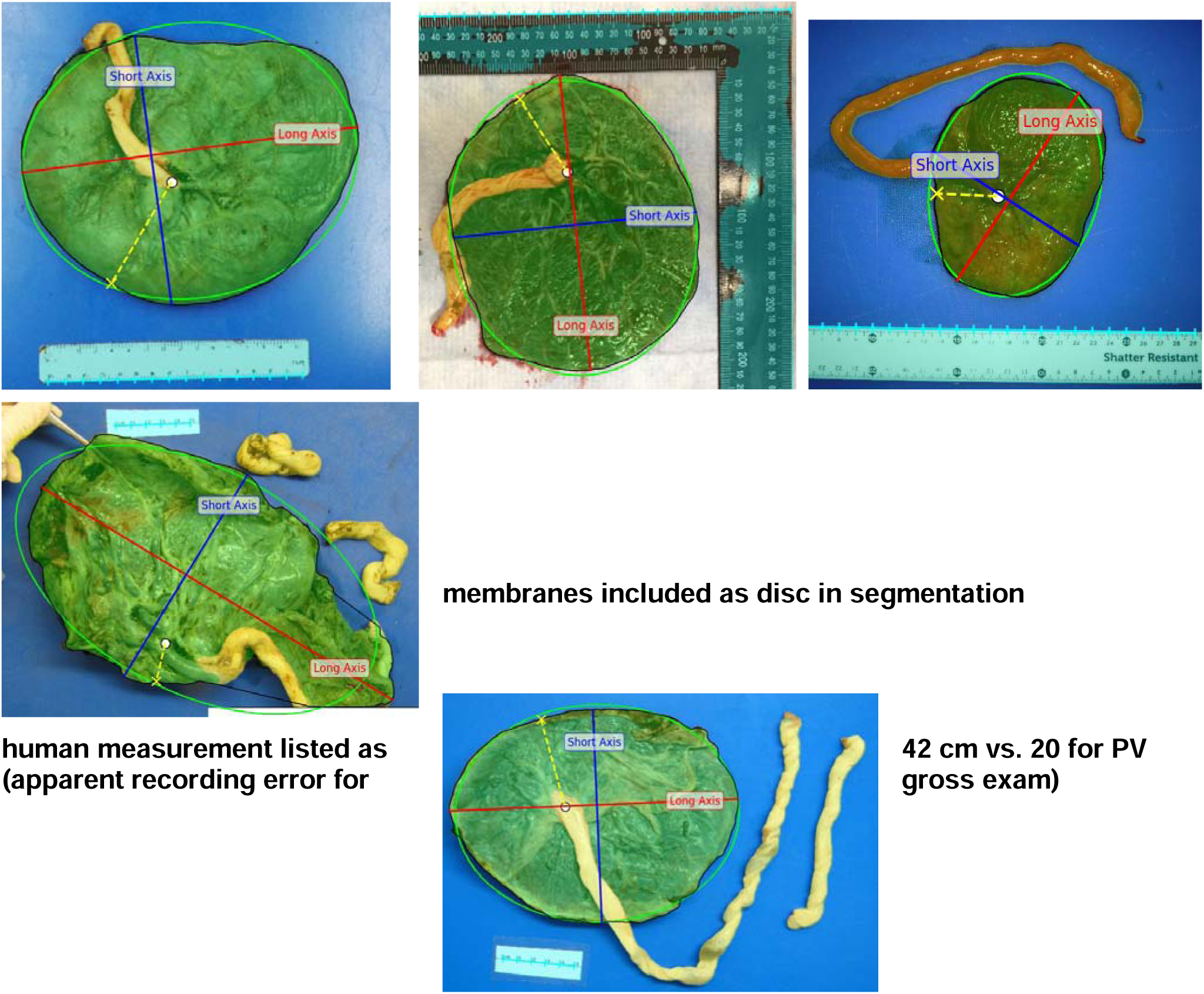
Example results for (row 1) good agreement, (row 2) PlacentaVision error (longer than gross exam), and (row 3) Gross exam error (longer than PlacentaVision)

**Supplemental Table 1.** Placental area and perimeter from PlacentaVision models, overall and by placenta shape, site, infant sex, and gestational age, n=27,846

|  | Area (cm<sup>2</sup>) |  | Perimeter (cm) |  |
| --- | --- | --- | --- | --- |
|  | Mean | SD | Mean | SD |
| Overall | 234.78 | 64.88 | 59.36 | 8.75 |
| Site |  |  |  |  |
| Northwestern | 229.99 | 62.55 | 58.90 | 8.75 |
| Magee | 275.48 | 60.42 | 63.90 | 7.38 |
| MUST | 275.95 | 75.65 | 62.78 | 7.97 |
| Shape<sup>2</sup> |  |  |  |  |
| Regular shape | 232.93 | 63.18 | 58.62 | 8.06 |
| Irregular shape<sup>2</sup> | 249.92 | 75.73 | 65.43 | 11.39 |
| Infant sex |  |  |  |  |
| Male<sup>3</sup> | 236.74 | 64.69 | 59.57 | 8.68 |
| Female | 234.59 | 63.53 | 59.39 | 8.55 |
| Gestational age at birth<sup>4</sup> |  |  |  |  |
| Term | 244.38 | 62.75 | 60.60 | 8.35 |
| Preterm<sup>4</sup> | 202.25 | 61.31 | 55.15 | 8.76 |

<sup>1</sup> Measured longest length through center of disc; measured width perpendicular to length through center of disc.
<sup>2</sup> defined per ellipse ratio measured by PlacentaVision
<sup>3</sup> medical record categories included male, female, and U; U=178 (not included in results by infant sex)
<sup>4</sup> Term ≥37 weeks gestation; preterm <37 weeks gestation

